# Sex differences in atrial myocardial fibrosis and degeneration: evaluation using left atrial low-voltage areas during catheter ablation of atrial fibrillation

**DOI:** 10.1101/2023.02.20.23286210

**Authors:** Masaharu Masuda, Yasuhiro Matsuda, Hiroyuki Uematsu, Ayako Sugino, Hirotaka Ooka, Satoshi Kudo, Subaru Fujii, Mitsutoshi Asai, Osamu Iida, Shin Okamoto, Takayuki Ishihara, Kiyonori Nanto, Takuya Tsujimura, Yosuke Hata, Taku Toyoshima, Naoko Higashino, Sho Nakao, Toshiaki Mano

## Abstract

**Background:** Atrial myocardial degeneration predisposes to atrial fibrillation (AF), ischemic stroke, and heart failure. Studies suggest the presence of sex differences in atrial myocardial degeneration. This study aimed to delineate sex differences in the prevalence, predictors, and prognostic impact of left atrial low-voltage areas (LVAs).

**Methods:** This observational study included 1,488 consecutive patients undergoing initial ablation for AF. Voltage mapping was performed after pulmonary vein isolation during sinus rhythm. LVAs were defined as regions where bipolar peak-to-peak voltage was < 0.50 mV. Results: LVA prevalence was higher in women (37.6%) than in men (16.0%). High age, persistent form of AF, diabetes mellitus, and a large left atrium were shown to be common predictors in both sex categories. Heart failure and history of stroke/thromboembolic events were men-specific predictors of LVA existence. Women experienced more AF recurrence than men (31.1% vs. 25.7%, p = 0.027). LVA existence was significantly associated with increased AF recurrence in each sex category, with a respective hazard ratio, 95% confidence interval and p value of 2.45, 1.87-3.22, and < 0.0001 in men and 1.82, 1.33-2.49, and < 0.0001 in women.

**Conclusions:** LVA was more frequent in women than men, and predicted frequent AF recurrence irrespective of sex category.

## Introduction

In a majority of patients with atrial fibrillation (AF), the disease is thought to develop from an underlying diseased atrial myocardium.^1,2,3^ Left atrial bipolar voltage decreases with the progression of myocardial degeneration, including fibrosis and other pathophysiological changes.^4^ In the clinical setting, atrial voltage mapping is often performed to predict the probability of AF recurrence after catheter ablation^5,6^ and to guide ablation strategies.^5,7,8^ The presence of low-voltage area (LVA) at a certain region of the left atrium is reported to indicate a process of ongoing whole atrial myocardial degeneration.^4^

Sex differences in various aspects of cardiovascular disease have been highlighted.^9^ Some studies have reported that women are more likely to experience left atrial LVAs than men,^6,8^ suggesting that AF in women may derive from more advanced atrial myocardial degeneration than AF in men. To date, however, no detailed information regarding sex differences in LVAs has yet appeared.

Here, we aimed to delineate the sex differences in LVAs, including their prevalence, predictors, and prognostic impact.

## Methods

### Patients

This retrospective prospective observational study enrolled 1,488 consecutive patients who underwent the initial ablation of AF at Kansai Rosai Hospital from December 2014 to March 2022. Patients in whom the left atrial voltage map was not completed were excluded. Other exclusion criteria were age < 20 years, prior left atrial catheter or ablation, and prior MAZE surgery. The total study population of 1,488 patients was divided into 2 sex categories, with 987 men and 501 women. This study complied with the Declaration of Helsinki. Written informed consent for the ablation and participation in the study was obtained from all patients, and the protocol was approved by our institutional review board.

### Catheter ablation and voltage mapping

Pulmonary vein isolation was performed in all patients using a linear radiofrequency catheter or cryoballoon. Other ablations, including LVA homogenization, linear ablation, ablation for induced atrial tachycardias, and ablation for non-pulmonary-vein AF triggers were added at the discretion of the attending physicians. An electroanatomical mapping system (CARTO 3, [Biosense Webster, Inc., Diamond Bar CA, USA]; Ensite NavX, [Abbott, Abbott Park IL, USA]; or Rhythmia, [Boston Scientific, Boston MA, USA]) was used for ablation guidance and mapping.

Following pulmonary vein isolation, left atrial voltage mapping was performed under sinus rhythm or atrial paced rhythm from the right atrium. The multi-electrode mapping catheter used in most cases was a PENTARAY (Biosense webstar), Lasso NaV (Biosense webstar), HD grid (Abbott), or Orion (Boston Scientific). A 3.5- or 4.0-mm ablation catheter was used as an auxiliary in some areas with difficulty in catheter-tissue contact. The band pass filter was set at 30 to 500 Hz. Adequate endocardial contact was confirmed by distance to the geometry surface and stable electrograms. Mapping points were acquired to fill all color gaps on the voltage map. Respective fill and color interpolation thresholds were 15 mm and 23 mm for CARTO 3 and 20 mm and 7 mm for Ensite NavX. The interpolation threshold for Rhythmia was set at 5 mm. The presence of LVAs was determined when the size of total areas with a bipolar peak-to-peak voltage < 0.50 mV was ≥ 5 cm^2^.^10^

### Follow-up

Patients were followed every 4–8 weeks at the dedicated arrhythmia clinic of our institution for a minimum of 2 years. Routine ECGs were obtained at each outpatient visit, and 24-h ambulatory Holter monitoring was performed at 6 and 12 months post-ablation. When patients experienced symptoms suggestive of an arrhythmia, a surface ECG, ambulatory ECG, and/or cardiac event recording were also obtained. Either of the following events following the initial 3 months after ablation (blanking period) was considered to indicate AF recurrence: (1) atrial tachyarrhythmia (AF and regular atrial tachycardia) recorded on a routine or symptom-triggered ECG during an outpatient visit; or (2) atrial tachyarrhythmia of at least 30-s duration on ambulatory ECG monitoring. No antiarrhythmic drugs were prescribed after the ablation procedure unless AF recurrence was observed.

### Statistical analysis

Continuous data are expressed as the mean ± standard deviation or median (interquartile range). Categorical data are presented as absolute values and percentages. Tests for significance were conducted using the unpaired *t*-test, or nonparametric test (Mann-Whiney *U*-test) for continuous variables, and the chi-squared test or Fisher’s exact test for categorical variables. A logistic regression model was used to identify clinical factors associated with LVA existence. Cox proportional hazard models were used to determine the prognostic significance of LVA in the prediction of AF recurrence. Variables with a P value ≤ 0.05 in the univariate models were included in the multivariate analysis. AF recurrence-free survival rates were calculated using the Kaplan-Meier method. Survival curves between groups were compared with a 2-sided Mantel-Haenszel (log-rank) test. All analyses were performed using commercial software (SPSS version 26.0^®^, SPSS, Inc., Chicago IL, USA).

## Results

### Patient characteristics

Patient characteristics are compared between men and women in Table 1. Women were older, smaller in height, lighter in weight, thinner, more likely to have paroxysmal AF, and had a greater decrease in renal function and a higher CHA2DS2VASc score. Women also had higher clinical scores predicting the presence of LVAs (SPEED and DR-FLASH).^11,12^ On echocardiography, women patients had smaller atrial and ventricular chamber sizes, a higher left ventricular ejection fraction, and lower left ventricular mass.

**Table 1.** Baseline characteristics.

|  | **Men**<br>*n* = 987 | **Women**<br>*n* = 501 | *p* |
| --- | --- | --- | --- |
| **Age, years** | 66.5 ± 10.6 | 71.3 ± 8.2 | <0.0001 |
| **Height, cm** | 168.8 ± 6.7 | 153.6 ± 6.4 | <0.0001 |
| **Weight, Kg** | 70.0 ± 12.2 | 55.4 ± 10.9 | <0.0001 |
| **Body mass index, kg/m<sup>2</sup>** | 24.3 ± 3.7 | 23.5 ± 4.6 | <0.0001 |
| **AF type** |  |  | <0.0001 |
| *Paroxysmal, n (%)* | 335 (34) | 248 (50) |  |
| *Persistent AF lasting < 1 year, n (%)* | 557 (56) | 225 (45) |  |
| *Persistent AF lasting ≥ 1 year, n (%)* | 95 (10) | 28 (6) |  |
| **Hypertension, n (%)** | 560 (57) | 274 (55) | 0.45 |
| **Diabetes mellitus, n (%)** | 187 (19) | 82 (16) | 0.22 |
| **Heart failure, n (%)** | 208 (21) | 114 (23) | 0.46 |
| **Stroke or systemic thromboembolism, n (%)** | 77 (8) | 40 (8) | 0.90 |
| **Vascular disease, n (%)** | 106 (11) | 28 (6) | 0.001 |
| **CHA<sub>2</sub>DS<sub>2</sub>-VASc score** | 2.1 ± 1.4 | 3.4 ± 1.3 | <0.0001 |
| **eGFR, mL/min/1.73m<sup>2</sup>** | 63.5 ± 18.3 | 60.8 ± 19.0 | 0.008 |
| **NT-pro BNP, pg/ml** | 540 (174, 976) | 562 (142, 1266) | 0.55 |
| **BNP, pg/ml** | 106 (40, 207) | 126 (56, 278) | 0.13 |
| **DR-FLASH score** | 3.3 ± 1.2 | 4.2 ± 1.1 | <0.0001 |
| **SPEED score** | 1.9 ± 1.1 | 2.9 ± 1.1 | <0.0001 |
| **Echocardiography findings** |  |  |  |
| *Left atrial diameter, mm* | 41.2 ± 6.9 | 39.4 ± 6.6 | <0.0001 |
| *Left ventricular diastolic diameter, mm* | 48.0 ± 6.1 | 44.0 ± 5.4 | <0.0001 |
| *Left ventricular ejection fraction, %* | 60.0 ± 12.6 | 63.6 ± 11.1 | <0.0001 |
| *Left ventricular mass index, g/m<sup>2</sup>* | 108 ± 32 | 103 ± 31 | 0.003 |

AF, atrial fibrillation; GFR, glomerular filtration rate; NT-pro BNP, N-terminal prohormone of brain natriuretic peptide; BNP, brain natriuretic peptide.

**Table 2.** Factors associated with LVA existence among men.

|  | **LVA** |  | **Univariate** |  |  | **Multivariate** |  |  |
| --- | --- | --- | --- | --- | --- | --- | --- | --- |
|  | *With*<br>*n = 158* | *Without*<br>*n = 829* | *HR* | *95% CI* | *p* | *HR* | *95% CI* | *p* |
| **Age, years** | 72.9 ± 7.8 | 65.3 ± 10.7 | 1.09 | 1.07-1.12 | <0.0001 | 1.13 | 1.09-1.18 | <0.0001 |
| **Body mass index** | 23.9 ± 3.7 | 24.4 ± 3.6 | 0.96 | 0.91-1.01 | 0.088 |  |  |  |
| **Persistent AF, n (%)** | 126 (80) | 526 (63) | 2.27 | 1.50-3.43 | <0.0001 | 2.01 | 1.19-3.37 | 0.009 |
| **Hypertension, n (%)** | 86 (54) | 474 (57) | 0.90 | 0.64-1.26 | 0.52 |  |  |  |
| **Diabetes mellitus, n (%)** | 42 (27) | 145 (18) | 1.71 | 1.15-2.54 | 0.008 | 2.04 | 1.17-3.54 | 0.012 |
| **Heart failure, n (%)** | 51 (32) | 157 (19) | 2.04 | 1.40-2.97 | <0.0001 | 2.16 | 1.27-3.67 | 0.005 |
| **Stroke/thromboembolism, n (%)** | 19 (12) | 58 (7) | 1.82 | 1.05-3.15 | 0.033 | 2.37 | 1.04-5.39 | 0.039 |
| **Vascular disease, n (%)** | 24 (15) | 82 (10) | 1.63 | 0.999-2.67 | 0.051 |  |  |  |
| **Elevated BNP\*, n (%)** | 125 (81) | 470 (58) | 3.01 | 1.97-4.58 | <0.0001 | 1.41 | 0.84-2.35 | 0.19 |
| **eGFR, mL/min/1.73m<sup>2</sup>** | 59 ± 14 | 60 ± 12 | 0.98 | 0.98-0.993 | 0.001 | 1.001 | 0.990-1.01 | 0.83 |
| **Left atrial diameter, mm** | 43.1 ± 6.6 | 40.8 ± 6.8 | 1.05 | 1.02-1.08 | <0.0001 | 1.03 | 0.999-1.06 | 0.054 |
| **Left ventricular ejection fraction, %** | 58.5 ± 14.0 | 60.2 ± 12.3 | 0.989 | 0.98-1.002 | 0.11 |  |  |  |

Factors with $p < 0.05$ in the univariate analysis were incorporated in the multivariate analysis. \*Elevated BNP was defined as brain natriuretic peptide $> 100$ pg/ml or N-terminal pro brain natriuretic peptide $> 400$ pg/ml. AF, atrial fibrillation; GFR, glomerular filtration rate; LVA, low-voltage areas; HR, hazard ratio; CI, confidence interval.

**Table 3.**
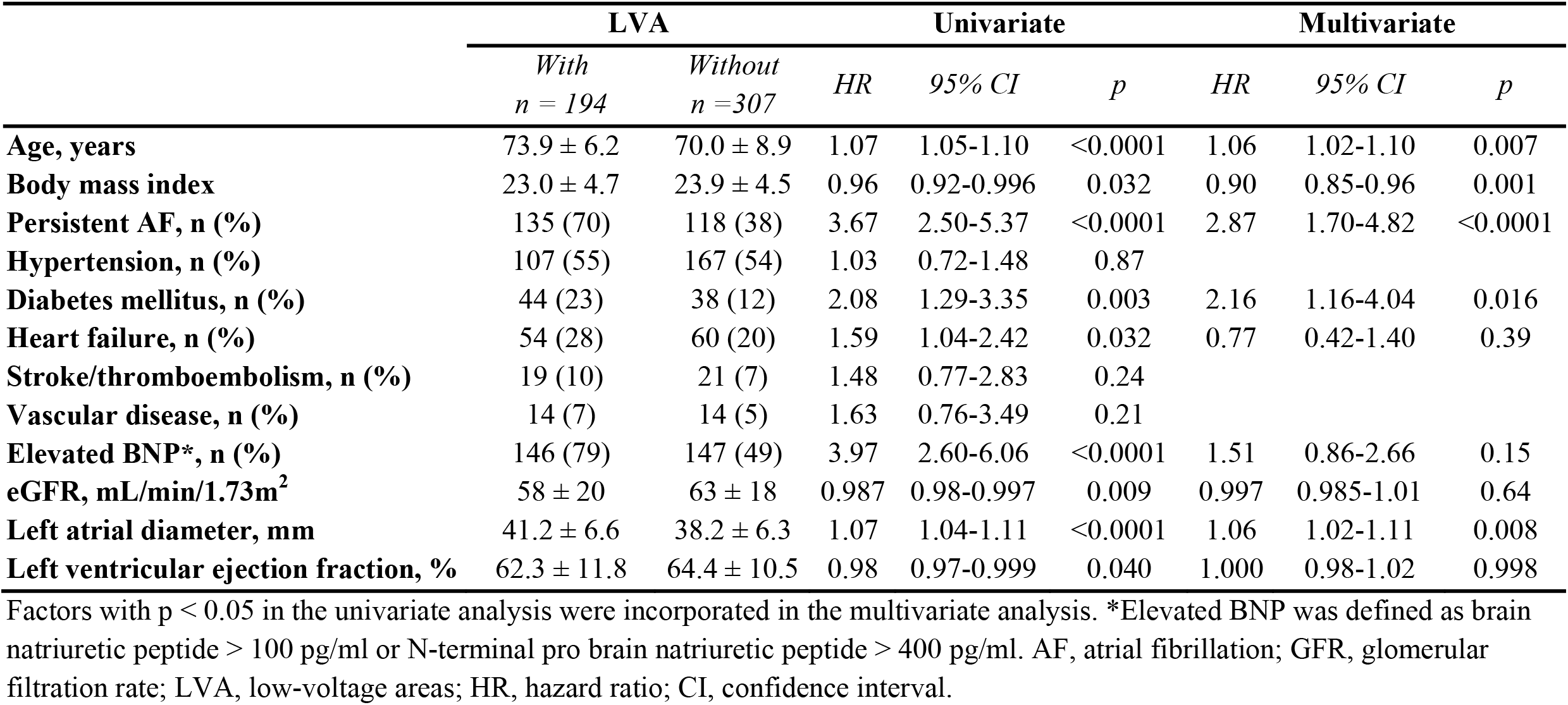
Factors associated with LVA existence among women.

|  | **LVA** |  | **Univariate** |  |  | **Multivariate** |  |  |
| --- | --- | --- | --- | --- | --- | --- | --- | --- |
|  | *With*<br>*n = 194* | *Without*<br>*n = 307* | *HR* | *95% CI* | *p* | *HR* | *95% CI* | *p* |
| **Age, years** | 73.9 ± 6.2 | 70.0 ± 8.9 | 1.07 | 1.05-1.10 | <0.0001 | 1.06 | 1.02-1.10 | 0.007 |
| **Body mass index** | 23.0 ± 4.7 | 23.9 ± 4.5 | 0.96 | 0.92-0.996 | 0.032 | 0.90 | 0.85-0.96 | 0.001 |
| **Persistent AF, n (%)** | 135 (70) | 118 (38) | 3.67 | 2.50-5.37 | <0.0001 | 2.87 | 1.70-4.82 | <0.0001 |
| **Hypertension, n (%)** | 107 (55) | 167 (54) | 1.03 | 0.72-1.48 | 0.87 |  |  |  |
| **Diabetes mellitus, n (%)** | 44 (23) | 38 (12) | 2.08 | 1.29-3.35 | 0.003 | 2.16 | 1.16-4.04 | 0.016 |
| **Heart failure, n (%)** | 54 (28) | 60 (20) | 1.59 | 1.04-2.42 | 0.032 | 0.77 | 0.42-1.40 | 0.39 |
| **Stroke/thromboembolism, n (%)** | 19 (10) | 21 (7) | 1.48 | 0.77-2.83 | 0.24 |  |  |  |
| **Vascular disease, n (%)** | 14 (7) | 14 (5) | 1.63 | 0.76-3.49 | 0.21 |  |  |  |
| **Elevated BNP\*, n (%)** | 146 (79) | 147 (49) | 3.97 | 2.60-6.06 | <0.0001 | 1.51 | 0.86-2.66 | 0.15 |
| **eGFR, mL/min/1.73m<sup>2</sup>** | 58 ± 20 | 63 ± 18 | 0.987 | 0.98-0.997 | 0.009 | 0.997 | 0.985-1.01 | 0.64 |
| **Left atrial diameter, mm** | 41.2 ± 6.6 | 38.2 ± 6.3 | 1.07 | 1.04-1.11 | <0.0001 | 1.06 | 1.02-1.11 | 0.008 |
| **Left ventricular ejection fraction, %** | 62.3 ± 11.8 | 64.4 ± 10.5 | 0.98 | 0.97-0.999 | 0.040 | 1.000 | 0.98-1.02 | 0.998 |

Factors with $p < 0.05$ in the univariate analysis were incorporated in the multivariate analysis. \*Elevated BNP was defined as brain natriuretic peptide $> 100$ pg/ml or N-terminal pro brain natriuretic peptide $> 400$ pg/ml. AF, atrial fibrillation; GFR, glomerular filtration rate; LVA, low-voltage areas; HR, hazard ratio; CI, confidence interval.

The specific three-dimensional mapping system used for ablation also differed between men and women, namely CARTO (81.3% in men vs. 77.8% in women), EnSite (9.6% vs. 13.8%), and Rhythmia (3.5% vs. 2.0%, p = 0.034). A cryoballoon was more frequently used for women than men (18.4% in men vs. 27.5% in women, p < 0.0001). Non-pulmonary-vein trigger ablation (3.1% in men vs. 5.6% in women, p = 0.026) and isolation of the superior vena cava (1.2% vs. 2.8%, p = 0.030) were more often performed in women than in men. Women more often underwent ablation targeting LVAs or areas with fractionated electrograms during sinus rhythm (5.9% in men vs. 13.0% in women, p < 0.0001). The use of other ablation procedures was similar in men and women, as follows: pulmonary vein isolation, 100% in men vs. 100% in women, p = 1.0; left atrial roof line, 6.3% vs. 7.9%, p = 0.31; left atrial bottom line, 1.2% vs. %, p = 0.073; mitral isthmus line, 1.0% vs. 0.6%, p = 0.76; and cavo-tricuspid isthmus line, 16.1% vs. 15.1%, p = 0.64.

### LVA prevalence

Left atrial LVA prevalence was higher in women (37.6%) than in men (16.0%, Figure 1). Prevalence of extensive LVA covering > 20 cm^2^ was also higher in women (12.2%) than in men (4.2%). In both sexes, LVAs were predominantly observed at the anterior-septal region, and higher LVA prevalence in women was consistently observed throughout every left atrial region.

**Figure 1.**
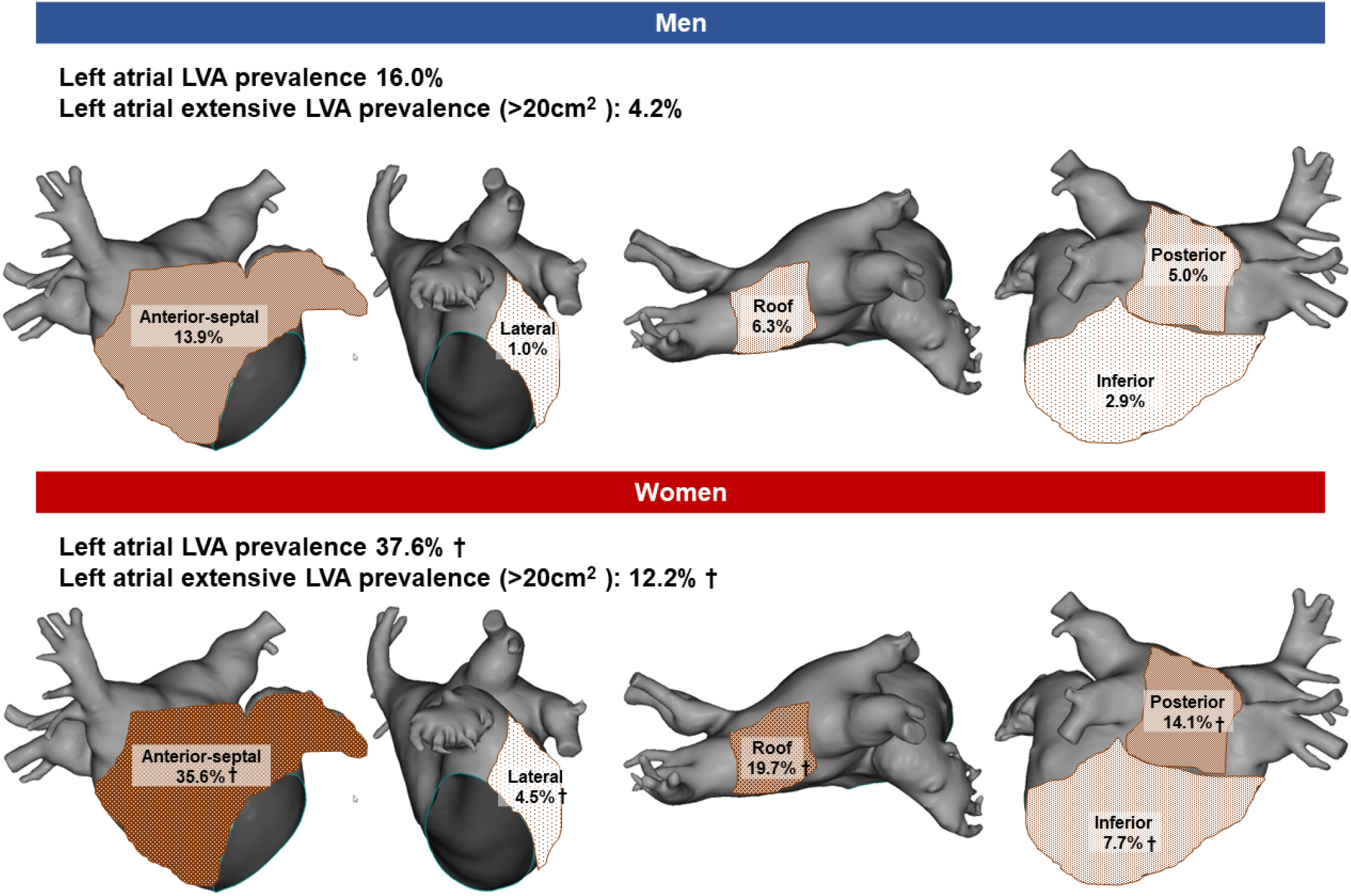
LVA prevalence and distribution. Prevalence of LVAs and extensive LVAs covering > 20 cm^2^, and distribution of LVAs are shown. LVA and extensive LVAs were more often observed in women than in men in whole left atrium and in each region. LVA indicates low-voltage area. †p< 0.05 for comparison between men and women.

Stratification by age group revealed a consistently higher LVA prevalence in women aged 60-84 years (Figure 2). No women aged ≤ 54 years old had LVAs. Figure 3 shows LVA prevalence stratified by left atrial diameter group. A higher LVA prevalence in women than men was consistently observed when left atrial diameter was ≥ 34 mm.

**Figure 2.**
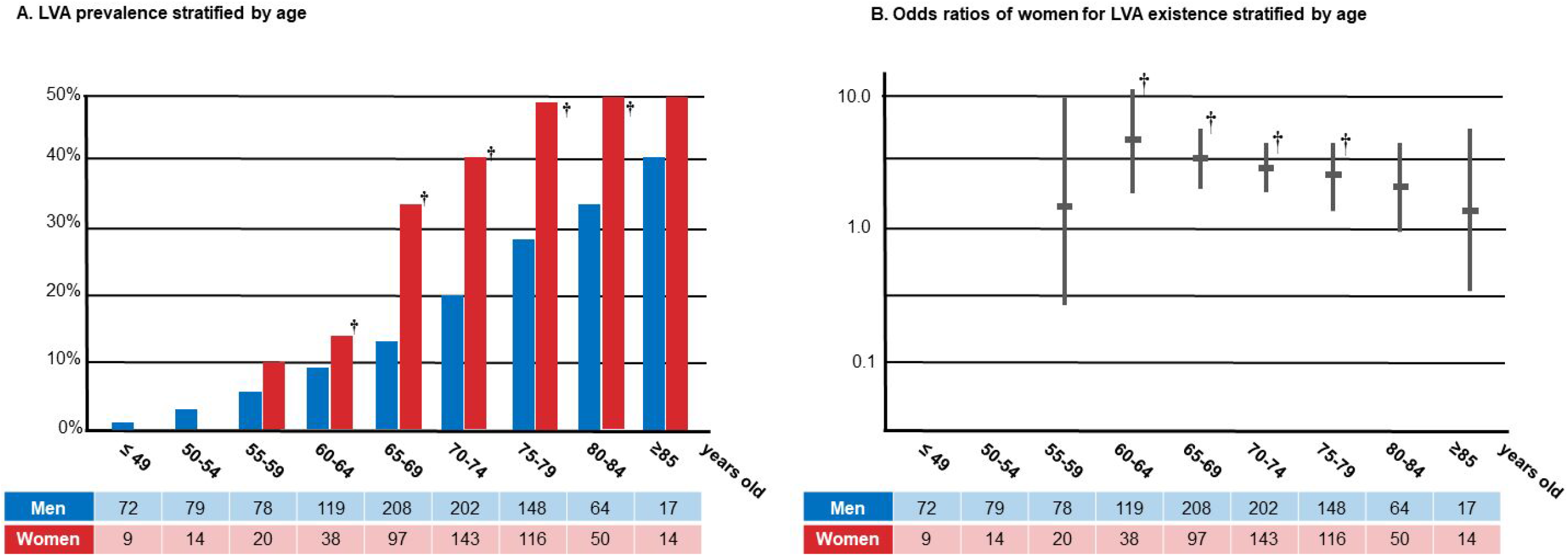
Age-stratified LVA prevalence and odds ratio of women for LVA existence. LVA prevalence (A) and odds ratios of women for LVA existence are shown stratified by age. A higher LVA prevalence in women than men was consistently observed in those aged 60-84 years old. No women aged ≤ 54 had LVAs. LVA indicates low-voltage areas. †p< 0.05 for comparison between men and women.

**Figure 3.**
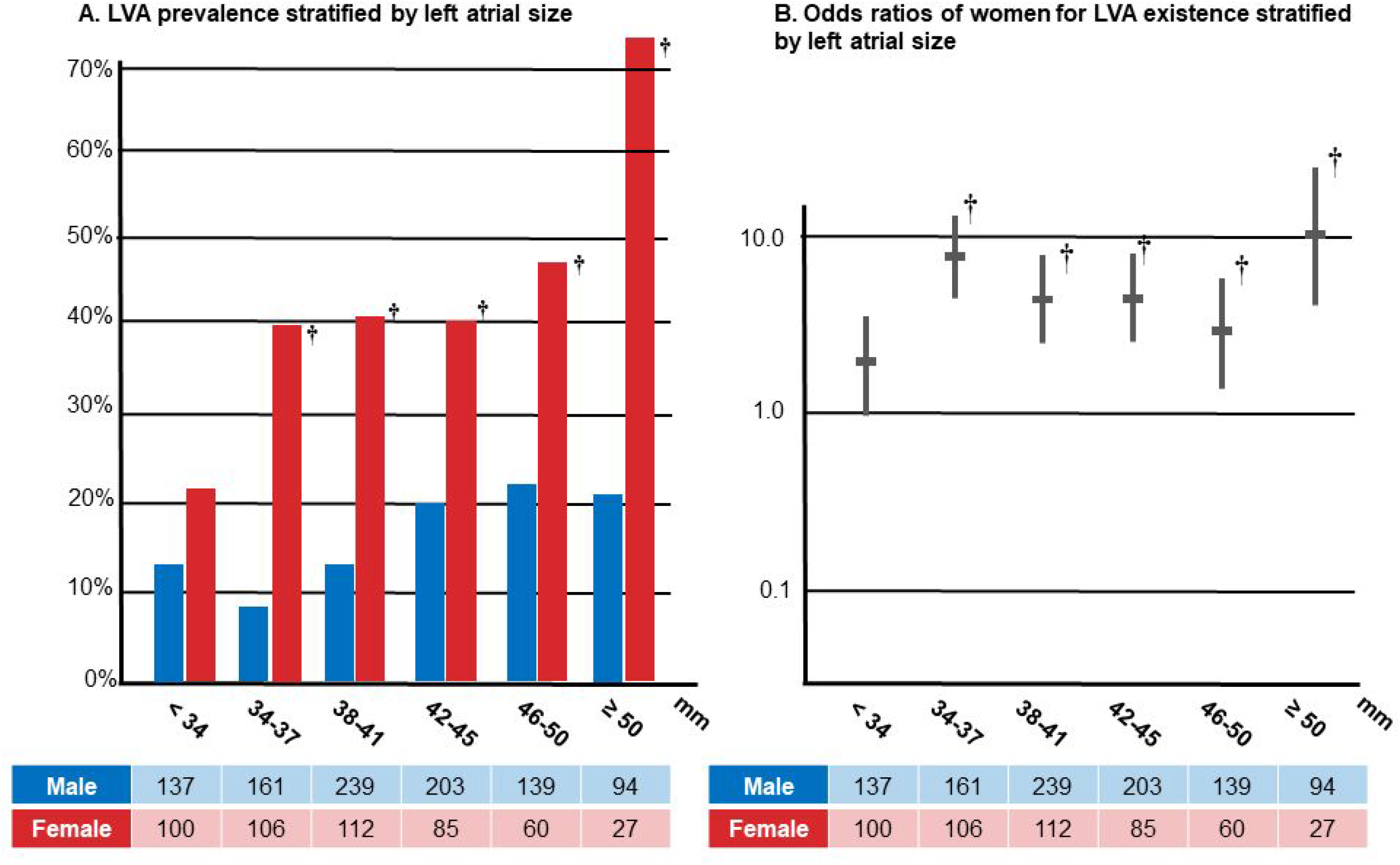
Atrial-size-stratified LVA prevalence and odds ratio of women for LVA existence. LVA prevalence (A) and odds ratios of women for LVA existence are shown stratified by left atrial size. A higher LVA prevalence in women than men was consistently observed when left atrial diameter was ≥ 34 mm. LVA indicates low-voltage areas. †p< 0.05 for comparison between men and women.

### Predictors of LVA existence in men and women

The association between LVA existence and clinical factors was investigated. High age, persistent form of AF, diabetes mellitus, and large left atrium were found to be common predictors in both sex categories. Heart failure and history of stroke/thromboembolic events were men-specific predictors of LVA existence.

### Prognostic significance of LVA existence in men and women

During a median follow-up period of 523 (207, 720) months, AF recurrence occurred in 410 of 1,488 (27.6%) patients. Kaplan-Meier analysis revealed that women experienced more AF recurrence than men (Figure 4). LVA existence was significantly associated with increased AF recurrence in both sex categories. LVA existence predicted AF recurrence after ablation with a respective hazard ratio, 95% confidence interval and p value of 2.45, 1.87-3.22, and < 0.0001 for men, and 1.82, 1.33-2.49, and < 0.0001 for women.

**Figure 4.**
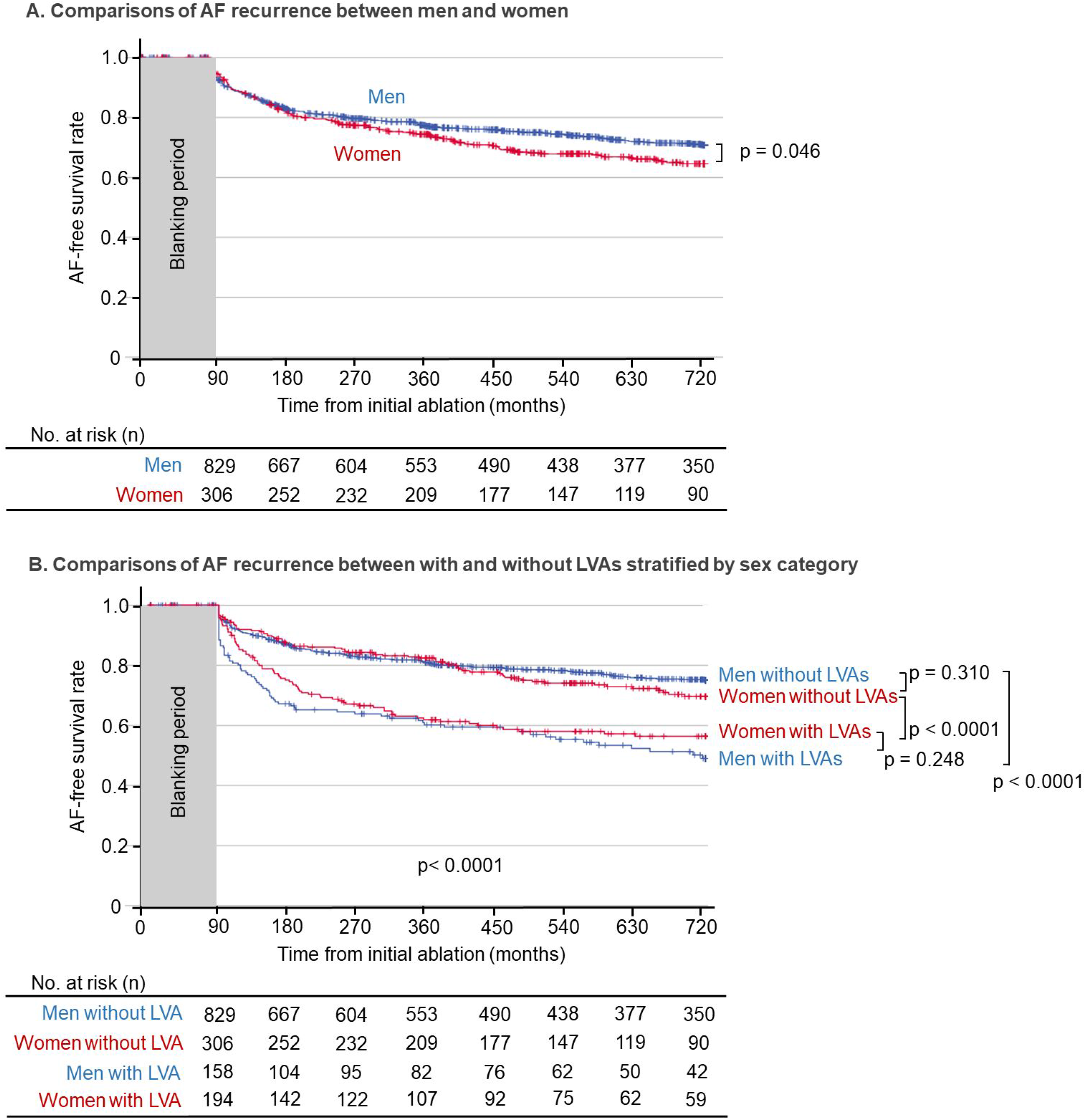
Kaplan-Meier curves of AF recurrence-free rates. Kaplan-Meier curves of AF recurrence-free rate after initial ablation. Comparison was performed between men and women (A), and 4 groups divided by sex category and LVA existence (B). Women experienced more AF recurrence than men. LVA existence was significantly associated with increased AF recurrence in each sex category.

## Discussion

This single-center retrospective study investigated sex differences in the prevalence, associated factors, and prognostic significance of LVAs. The main findings were as follows. First, LVA prevalence was higher in women than in men, irrespective of age and left atrial size. Second, LVAs were predominantly observed at the anterior-septal region in both men and women, with higher prevalence in women at each left atrial region. Third, common predictors of LVA existence were high age, persistent form of AF, diabetes mellitus, and large left atrium. Heart failure and a history of stroke were men-specific predictors of LVA existence. Finally, women developed AF recurrence more frequently, while LVA demonstrated prognostic significance regardless of sex. These findings delineate sex differences in LVAs and left atrial remodeling.

### Atrial myocardial degeneration in women

Reduction in atrial bipolar voltage is reported to be associated with atrial myocardial degeneration, as characterized by fibrosis, vacuolar degeneration, and deposition of abnormal substances such as amyloid-beta protein on histopathological analysis of atrial myocardial specimens obtained from the right atrial septum during the AF ablation procedure.^4^ On the other hand, several studies using magnetic-resonance cardiac imaging reported the coincidence of LVAs and areas with delayed gadolinium enhancement, suggesting that LVA reflects fibrosis.

The higher prevalence of LVAs in women, which has been previously reported,^12,13^ was confirmed across broad age and left atrial size groups. Several sex-specific pathophysiological features can be noted. For example, female are reported to have increased myocyte stiffness,^14^ and increased expression of connexin 40.^15^ Progesterone-induced activation of protein kinase B and extracellular signal-regulated kinase 1/2 could lead to pregnancy-induced cardiac hypertrophy.^16^ In addition, hormonal change during menopause influence myocardial remodeling in women, including a drop in nitric oxide with menopause,^17^ post-menopausal activation of the renin-angiotensin-aldosterone system in response to low estrogen,^17^ increased expression of protein kinase A.^18^ Our finding that LVAs in women were observed at age ≥55 years old and significantly increased at age ≥60 years old supports the possible influence of post-menopause-related mechanisms (Figure 2).

Another perspective also supports the possibility of a sex-specific difference in the underlying pathophysiology of AF development. Atrial remodeling can be classified into electrical and structural (anatomical and histological) remodeling, both of which likely contribute to causing electrical abnormalities such as reentry and ectopic firing, and AF.^19^ We hypothesize that AF development in women requires advanced atrial myocardial degeneration, whereas other factors such as electrical remodeling and anatomical enlargement play a predominant role in AF development in men. This hypothesis appears supported by the higher incidence of AF in men than women,^20^ despite women AF patients having more severe left atrial myocardial degeneration.

### Associated factors of atrial myocardial degeneration

We found that high age, persistent form of AF, and diabetes mellitus were common predictors of LVA existence, irrespective of sex. These factors have been repeatedly reported in prior studies, and are well known to be associated with atrial myocardial degeneration.^12,21-25^ Notably, a history of heart failure and stroke/thromboembolic events has been associated with LVA existence specifically in men. Elevated atrial pressure in heart failure patients can advance atrial remodeling, and lead to atrial myocardial degeneration and LVA existence. Left atrial thrombus is easier to generate on diseased atrial endocardium, and stroke and thromboembolic events are reportedly more frequent in patients with atrial cardiomyopathy.^1^ The impact of heart failure and stroke/thromboembolic events might be conspicuous in men because men are relatively less influenced by sex-specific mechanisms predisposing to atrial myocardial degeneration.

### AF recurrence after catheter ablation

Our finding that AF recurrence rate was higher in women than in men is consistent with prior studies.^26,27^ On the other hand, this sex difference disappeared on comparison among patients with and without LVAs (Figure 4), which in turn suggests that the sex difference in AF recurrence after ablation is due to a difference in the severity of atrial myocardial degeneration.

Atrial fibrotic degeneration is reported to lead to an atrial arrhythmogenic substrate that is associated with conduction disturbance and shortening of action potential duration, which in turn predisposes to non-pulmonary-vein AF trigger and maintenance of AF reentrant activity.^28-31^ Indeed, we found here that LVA existence was associated with frequent AF recurrence in both men and women. Considering that the majority of ablation procedures in this study population was pulmonary vein isolation alone, LVA existence indicated the presence of extra-pulmonary vein arrhythmogenic substrate.

### Clinical implications

The results of this study can be interpreted to mean that AF in women more frequently derives from myocardial degeneration. This would explain, partially at least, two major characteristics of AF in women, namely poor AF ablation outcomes and frequent ischemic stroke.^1,27^ Clinical studies designed to delineate sex differences are needed to improve sex-specific management of AF patients.

### Limitations

Several limitations of this study warrant mention. First, as we conducted voltage mapping after completion of PVI, LVAs near the pulmonary vein antrum could not be determined precisely due to ablation scar. Second, small LVAs (< 5cm^2^) were not considered diagnostic of LVA existence in order to avoid misjudgment. Third, voltage maps and LVAs were possibly influenced by different mapping catheters and different mapping systems. These factors can influence the results of a voltage map. Fourth, selection of ablation procedures was at the discretion of the operator, albeit based on international guidelines and electrophysiological findings, and were accordingly not necessarily consistent. This might have biased the AF recurrence rate. Fifth, AF recurrence after discharge was quantified on the basis of the patient’s symptoms, giving rise to the possibility that asymptomatic episodes of AF might have been missed.

### Conclusion

LVA prevalence was higher in women than in men, irrespective of age and left atrial size, suggesting that AF in women is more frequently derived from myocardial degeneration. Although LVA existence predicted AF recurrence, irrespective of sex category, the higher LVA prevalence in women led to the more frequent recurrence of AF in women. These results suggest the need to improve the sex-specific management of AF.

## Data Availability

Please contact the corresponding author by email

